# Self-reported pharmacogenetic medication use in the *Our Future Health* cohort

**DOI:** 10.1101/2025.09.03.25335018

**Authors:** Padraig Dixon, William G. Newman, Videha Sharma, John H McDermott, Cynthia Wright Drakesmith

**Affiliations:** Nuffield Department of Primary Care Health Sciences, University of Oxford, Oxford, UK; Manchester Centre for Genomic Medicine, St Mary’s Hospital, Manchester University NHS Foundation Trust, Manchester, UK; Division of Evolution and Genomic Sciences, School of Biological Sciences, University of Manchester, Manchester, UK

## Abstract

**Aim:** To describe self-reported use of medications with pharmacogenetic guidance in the Our Future Health (OFH) cohort.

**Methods:** We focused on four key pharmacogenes—*CYP2C19, CYP2C9, CYP2D6*, and *SLCO1B1*—and associated medications supported by high levels of evidence for clinical actionability according to the Clinical Pharmacogenetics Implementation Consortium (CPIC). We summarized self-reported pharmacogenetic medication use, assessed concurrent use, and stratified the findings by age, sex, and ethnicity.

**Results:** Self-reported medication use was analysed in 1.78 million OFH cohort participants included in the June 2025 data release. The cohort was 57.3% female, ranged in age from 18 to 95 years (mean 53.1 years), and 90.2% self-identified as being of “White” ethnicity. Participants reported use of 18 medication groups that were explicitly mentioned in the baseline questionnaire. Due to the structure of the baseline questionnaire, it was only possible to identify exposure to medication groups rather than mention of specific individual medications. Medications and medication groups with pharmacogenetic guidance included anti-depressants (selective serotonin reuptake inhibitors and tricyclics), statins, proton pump inhibitors, ibuprofen, opioids, clopidogrel and warfarin. A total of 25.2% (N=449,641) of the cohort reported using at least one of these medication groups, and were older, more likely to be female, and reported having more medical conditions than non-users. Concurrent use of these medications/medication groups metabolised by more than one of the four genes was common (37%).

**Conclusions:** Exposure to medications with pharmacogenetic guidance was prevalent, occurred in all age groups, and increased with age. The Our Future Health cohort will become an increasingly valuable resource for studying pharmacogenetics with further recruitment and the planned linkage to patient-level primary care prescribing data.

## INTRODUCTION

Each year, over one billion prescriptions are dispensed in England within the National Health Service (NHS) (1). However, variability in individual medication response remains a significant challenge (2, 3). Variation in genetic makeup plays a significant role; approximately 25% of all new primary care prescriptions involve drug–gene interactions. Furthermore, almost all individuals carry a genetic variant that may influence the safety or effectiveness of medication (5).

Pharmacogenetic (PGx) testing could support improved medication selection and dosing, reduce the risk of adverse drug reactions, and improve therapeutic outcomes by accounting for how genetic variation influences drug effectiveness and safety. However, despite this promise, PGx testing in routine care within the NHS remains limited in scope and application (6), although the most recent (July 2025) NHS 10-year plan proposes to integrate pharmacogenetics into routine clinical care (7). Given the widespread use of PGx-relevant medications among NHS patients, and the policy commitment to significantly expand pharmacogenetic testing, there is a clear need to better understand prescribing trends—particularly patterns of PGx medication use across patient demographics and in the context of polypharmacy.

This study aims to address these gaps by characterizing prescribing patterns of medications for which there are established pharmacogenomic prescribing guidelines using self-reported data on medication use from the Our Future Health (OFH) cohort (8). OFH is recruiting up to five million adults across the UK to support research on disease prevention, detection, and treatment. The OFH cohort will ultimately link genetic data, survey responses, and prescription records. However, at the time of writing, prescription data are not yet available; only self-reported medication use is accessible. In this study, we described self-reported medication use for which there is robust pharmacogenetic guidance (referred herein as PGx medications) in the most recently available data release.

## METHODS

### Overview

Baseline questionnaire data from the Our Future Health (OFH) cohort was used to assess self-reported use of PGx medications. The analysis focused on medications with the highest level of evidence for four pharmacogenes, as defined by the Clinical Pharmacogenetics Implementation Consortium (CPIC), and was based on the most recent data release of June 2025.

### OFH recruitment

The OFH cohort is a population-based, volunteer research programme open to adults (aged ≥18 years) living in the UK (8). The programme aims to recruit five million participants. Recruitment is ongoing and is primarily community-based, with the majority of participants joining through postal invitations. These were either personalised (via NHS DigiTrials) or non-personalised (to all adult residents within selected postcode areas) (8, 9). Other recruitment pathways included invitations to pharmacy customers and to registrants with the NHS Blood and Transplant service (8).

### Ethical approval

Ethical approval for Our Future Health was granted by the Cambridge East Research Ethics Committee. (REC reference: 21/EE/0016; date of opinion: 29 March 2021). The present study was approved on application to Our Future Health as project OFHS240243.

### Selection of pharmacogenetic drug-gene pairs

We focused on four pharmacogenes: *CYP2C19, CYP2D6, SLCO1B1*, and *CYP2C9*. Certain variants in these genes are associated with differential medication response. For example, Kimpton et al (10) reported that three of these genes (*CYP2D6, CYP2C19*, and *SLCO1B1*) influenced the metabolism of over 95% of all PGx drugs prescribed in a longitudinal study of NHS primary care prescribing.

To identify medications associated with the four target genes, we used classifications from the Clinical Pharmacogenetic Implementation Consortium (CPIC), which assigns evidence levels to drug–gene pairs based on the strength and quality of scientific and clinical data. Drug–gene pairs rated as Level A have the highest level of evidence and are characterised by CPIC as “likely to be clinically actionable”. We accessed the CPIC drug-gene pair list (11) on 30 May 2025, and identified candidate drugs and drug groups on a gene-by-gene basis. We selected only drugs with a “Final” rather than “Provisional” level status.

### Self-reporting of medication

Participants were asked to report medication use. One example that indicates the general structure of these questions is as follows: “Do you regularly take any of the following medication for heart or circulatory disorders (You can select more than one answer)” These questions were accompanied by the following statement “By regularly we mean something you take on a fixed schedule, or on most days of the week for the last four weeks. If you know you take a medication regularly, but don’t recognise these names, please check the packaging. Sometimes the brand names on the packaging will not be the same as the key ingredients.”

In turn, the options available were, in general, a mixture of single specific drugs (e.g. azathioprine), types of drugs without specific named examples (e.g. JAK inhibitors), or types of drugs with specific named examples such as “Proton pump inhibitors (e.g., omeprazole, esomeprazole, lansoprazole, rabeprazole, pantoprazole, dexlansoprazole)”. In other cases, the baseline questionnaire referred to types of medication (e.g. “Cholesterol lowering medication/statins”), but it appears participants were prompted with examples of specific statins when answering this question.

We assessed responses for specific mentions of individual medications or medication groups. After identifying all unique responses to these medication-related questions, we applied regular expression searches using exact word-level matching for generic names, brand names, and a limited set of known aliases. If a medication/medication group meeting CPIC drug-gene inclusion criteria was used but not explicitly named in the response, its use would not be detected and was therefore not included in our analysis. Otherwise, drugs explicitly mentioned in responses were taken as evidence of use. These limitations are important when interpreting our findings, and we address them further in the Discussion section. Additional details on classifications and questionnaire items contained in the baseline questionnaire are provided in Supplementary Material S2.

### Analysis

We summarized the cohort’s demographic characteristics—age, sex, ethnicity, household income, educational attainment and body mass index (BMI)—grouped by use of PGx medications. We described the use of specific medication categories (e.g., SSRIs, statins), patterns of PGx medication use by age, sex and ethnicity, and how many individuals used one or more PGx medication groups. Additionally, we reported how many participants used multiple PGx medication groups at the time of the baseline recruitment questionnaire and identified which of the four key genes were involved.

### Code and data availability

The code used to undertake this analysis is available from github.com/pdixon-econ/ofh_pgx. Access to the June 2025 data release was facilitated by the Our Future Health Early Adopters “Research Scholar” programme in which authors Dixon and Wright Drakesmith participated. The data were analysed in the Our Future Health DNAnexus Trusted Research Environment (https://ourfuturehealth.dnanexus.com/). Access to the data used in our analysis will be facilitated to qualifying researchers on application to Our Future Health.

## RESULTS

A total of 1,781,891 individuals had completed the baseline health questionnaire in the June 2025 data release. Responses were drawn from participants consented between 2021 and 2025, of which 83.5% were consented in 2023 and 2024.

### Identification of drug-gene pairs and associated medications

A total of 18 medications with CPIC Level A evidence associated with CYP2C19, CYP2D6, SLCO1B1, or CYP2C9 metabolizer status and with a finalized “Status Level”, were recorded in questionnaire responses and therefore included in our analysis. These 18 medications are: amitriptyline, atorvastatin, citalopram, clopidogrel, codeine, escitalopram, fluvastatin, ibuprofen, lansoprazole, omeprazole, pantoprazole, paroxetine, pravastatin, rosuvastatin, sertraline, simvastatin, tramadol, and warfarin. A further 16 medications met the CPIC Level A criteria for these four genes but were not explicitly mentioned. These medications, named in Supplementary Material S3, did not form part of our analysis.

A total of 449,641 participants (25.2%) reported the use of at least one pharmacogenetic (PGx) medication group from those explicitly mentioned, while 1,332,250 participants (74.8%) did not. The medication groups are statins, tricyclics, selective serotonin reuptake inhibitors (SSRIs), proton pump inhibitors (PPIs), non-steroidal anti inflammatory drugs, opioids, anticoagulants and antiplatelets. The number of individuals reporting PGx use by medication group, with specific medications explicitly mention in the baseline questionnaire, is shown in Figure 1.

**Figure 1.**
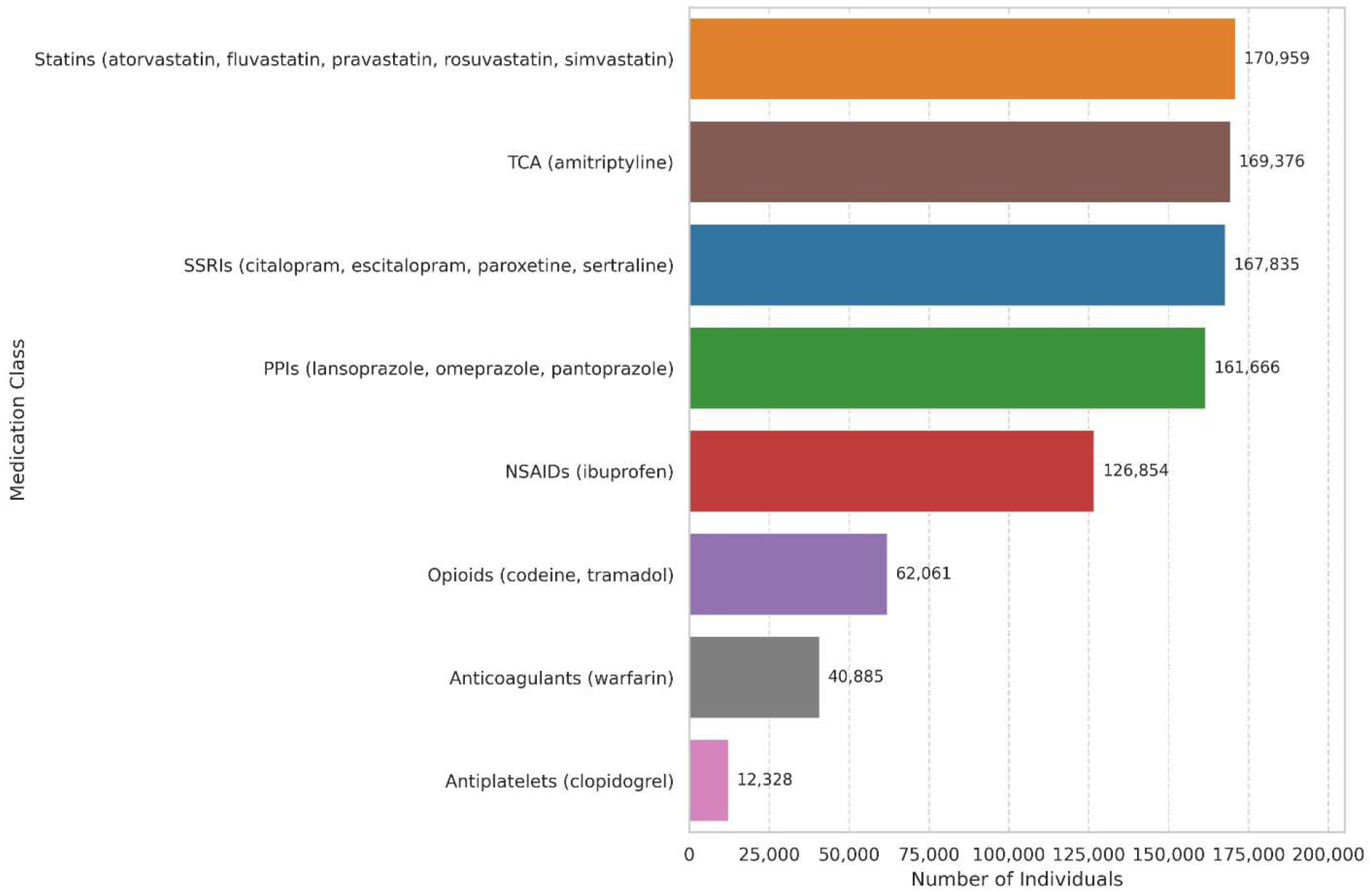
Self-reported use of medication groups

### Demographic information stratified by PGx medication use

Mean age at the time of consent was 53.1 years, and the cohort ranged in age from 18 to 95. The cohort was 57.3% female and was primarily of White ethnicity (Table 1). The percentage of “White British”, part of the underlying data used to create the overall “White” category, was 81.8% compared to 77.7% in the most recent census data (12).

**Table 1.** Characteristics of participants responding to specific baseline questionnaire items

| Characteristic | Statistics | All participants | Using named PGx medication / medication groups | Not using named PGx medication / medication groups |
| --- | --- | --- | --- | --- |
| **Sex (Female)** | N (%) | 1,020,110 (57.3%) | 281,284 (62.6%) | 738,826 (55.5%) |
| **Age (years)** | Mean (Standard deviation) | 53.1 (15.9) | 55.2 (16.0) | 52.4 (15.8) |
| **Age bands (years)** | N (%) |  |  |  |
| 18-30 |  | 168,480 (9.5%) | 36,876 (8.2%) | 131,604 (9.9%) |
| 31-45 |  | 405,505 (22.8%) | 87,232 (19.4%) | 318,273 (23.9%) |
| 46-60 |  | 520,517 (29.2%) | 128,394 (28.6%) | 392,123 (29.4%) |
| 61-75 |  | 554,533 (31.1%) | 151,487 (33.7%) | 403,046 (30.3%) |
| 76-90 |  | 131,232 (7.4%) | 44,973 (10.0%) | 86,259 (6.5%) |
| 90+ |  | 1,336 (0.1%) | 545 (0.1%) | 791 (0.1%) |
| **Ethnicity** | **N (%)** |  |  |  |
| White |  | 1,603,254 (90.2%) | 422,836 (94.2%) | 1,180,418 (88.8%) |
| Arab |  | 4,283 (0.2%) | 897 (0.2%) | 3,386 (0.3%) |
| Black |  | 36,533 (2.1%) | 6,687 (1.5%) | 29,846 (2.2%) |
| Chinese |  | 18,848 (1.1%) | 1,386 (0.3%) | 17,462 (1.3%) |
| Mixed |  | 9,561 (0.5%) | 2,234 (0.5%) | 7,327 (0.6%) |
| Other |  | 17,332 (0.9%) | 3,451 (0.7%) | 13,881 (1.0%) |
| South Asian |  | 88,441 (5.0%) | 11,524 (2.6%) | 76,917 (5.8%) |
| **BMI (kg/m<sup>2</sup>)** | **Mean**<br>**(Standard deviation)** | 26.7 (5.6) | 28.3 (6.4) | 26.2 (5.2) |
| **BMI categories** | **N (%)** |  |  |  |
| Underweight |  | 22,652 (1.4%) | 4,551 (1.1%) | 18,101 (1.5%) |
| Normal |  | 685,876 (42.0%) | 126,864 (31.2%) | 559,012 (45.6%) |
| Overweight |  | 581,295 (35.6%) | 147,650 (36.3%) | 433,645 (35.4%) |
| Obese |  | 343,627 (21.0%) | 127,726 (31.4%) | 215,901 (17.6%) |
| **Household income** | **N(%)** |  |  |  |
| < £18,000 |  | 169,606 (11.4%) | 64,975 (17.4%) | 104,631 (9.3%) |
| £18,000 - £30,999 |  | 271,545 (18.2%) | 83,236 (22.3%) | 188,309 (16.8%) |
| £31,000 - £51,999 |  | 366,048 (24.5%) | 94,732 (25.4%) | 271,316 (24.2%) |
| £52,000 - £100,000 |  | 462,024 (31.0%) | 95,992 (25.7%) | 366,032 (32.7%) |
| > £100,000 |  | 223,484 (15.0%) | 34,719 (9.3%) | 188,765 (16.9%) |
| **Educational qualifications** | **N(%)** |  |  |  |
| Vocational |  | 138,751 (8.2%) | 44,560 (10.6%) | 94,191 (7.4%) |
| Professional |  | 40,664 (2.4%) | 11,888 (2.8%) | 28,776 (2.3%) |
| Secondary education |  | 569,864 (33.6%) | 161,354 (38.5%) | 408,510 (32.0%) |
| Degree |  | 920,232 (54.3%) | 195,207 (46.5%) | 725,025 (56.8%) |
| Other |  | 26,620 (1.6%) | 6,386 (1.5%) | 20,234 (1.6%) |

Note: Numbers are those reporting responses in each category and are typically less than total number of participants in the overall cohort.

Participants differed demographically depending on whether they used the PGx medication/medication groups included in our analysis; in simple unadjusted two-sample t-tests, all reported comparisons differed with p<0.001. Among all participants, 57.3% were female (compared to 51.7% in the general population in the most recent UK census (12)) but females made up a higher proportion (62.6%) of those using PGx medications compared to 55.5% of those not using them. PGx medication/medication group users were also slightly older. Figure 2 shows the proportion of PGx use by age and sex, indicating greater use of PGx medication by increasing age band groups, and relatively higher use by males in older age bands. Note, however, that the absolute number of individuals in these older age bands (see Table 1) represents only a small proportion of the total cohort.

**Figure 2.**
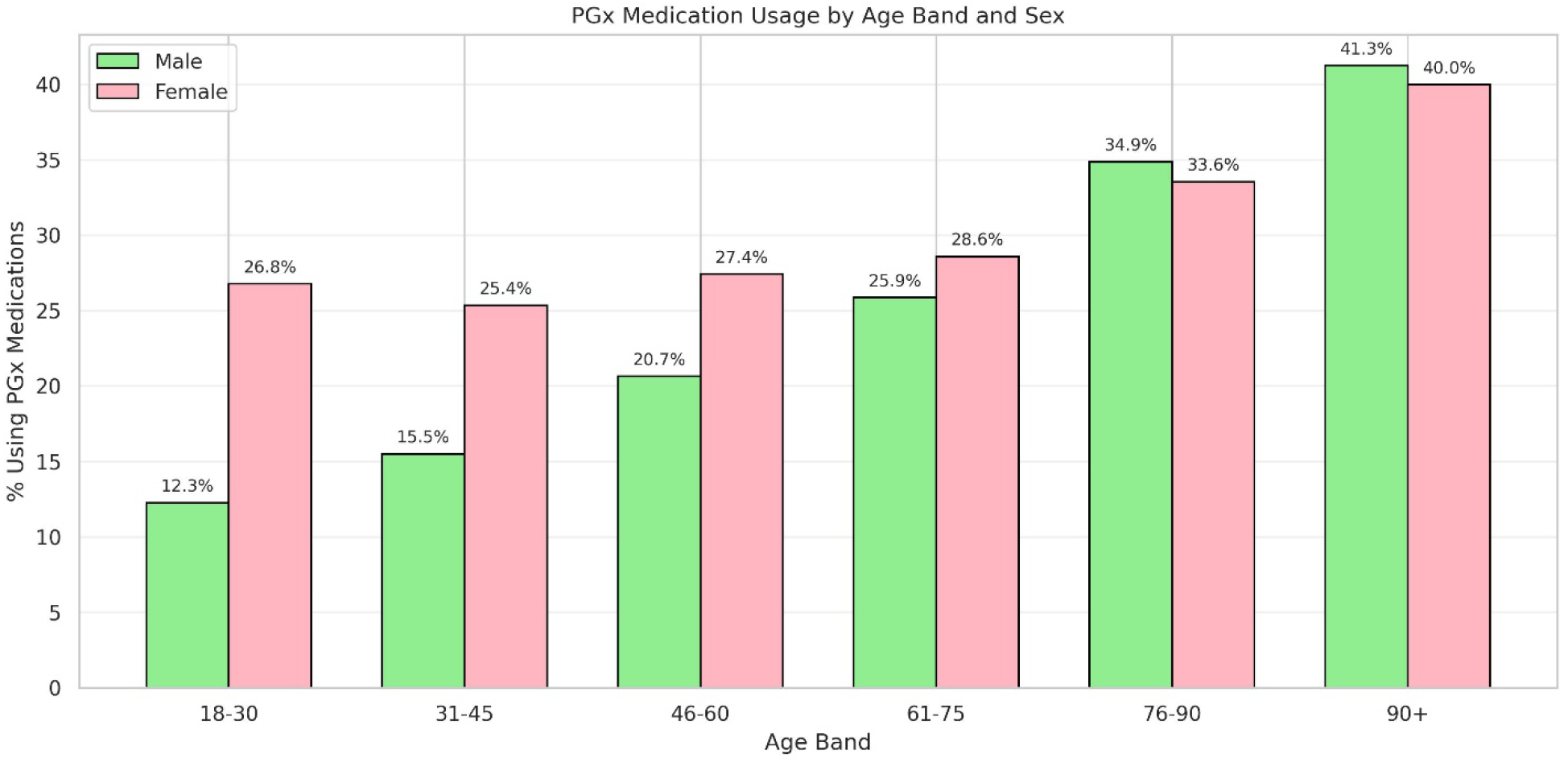
Self-reported PGx medication use by age band and sex

The PGx medication users had higher mean BMI (28.3 vs 26.2) and were more likely to be obese (31.4% vs 17.6%). PGx medication users’ mean BMI was slightly higher than mean adult BMI in recent estimates from the Health Survey for England (13) of 27.5kg/m^2^ and were more likely to be either overweight or obese (58% in that survey compared to 68% in the PGx medication group). PGx medication users were more likely to report lower household income (<£18,000: 17.2% vs. 9.3%), less likely to report household income >£100,000 (9.3% vs. 16.9%) and were less likely to have a degree.

### Self-reported health conditions stratified by PGx drug use

Generally, PGx users were more likely to report having received diagnoses of health conditions, the most prevalent of which were anxiety disorders, bone and joint conditions, depression, cardiovascular disease and gastrointestinal disorders (Figure 3). With the exception of bone and joint disorders, each of these conditions is relevant to CPIC guidelines pertaining to the four target genes regarding clinical actionability for PGx-informed prescribing.

**Figure 3.**
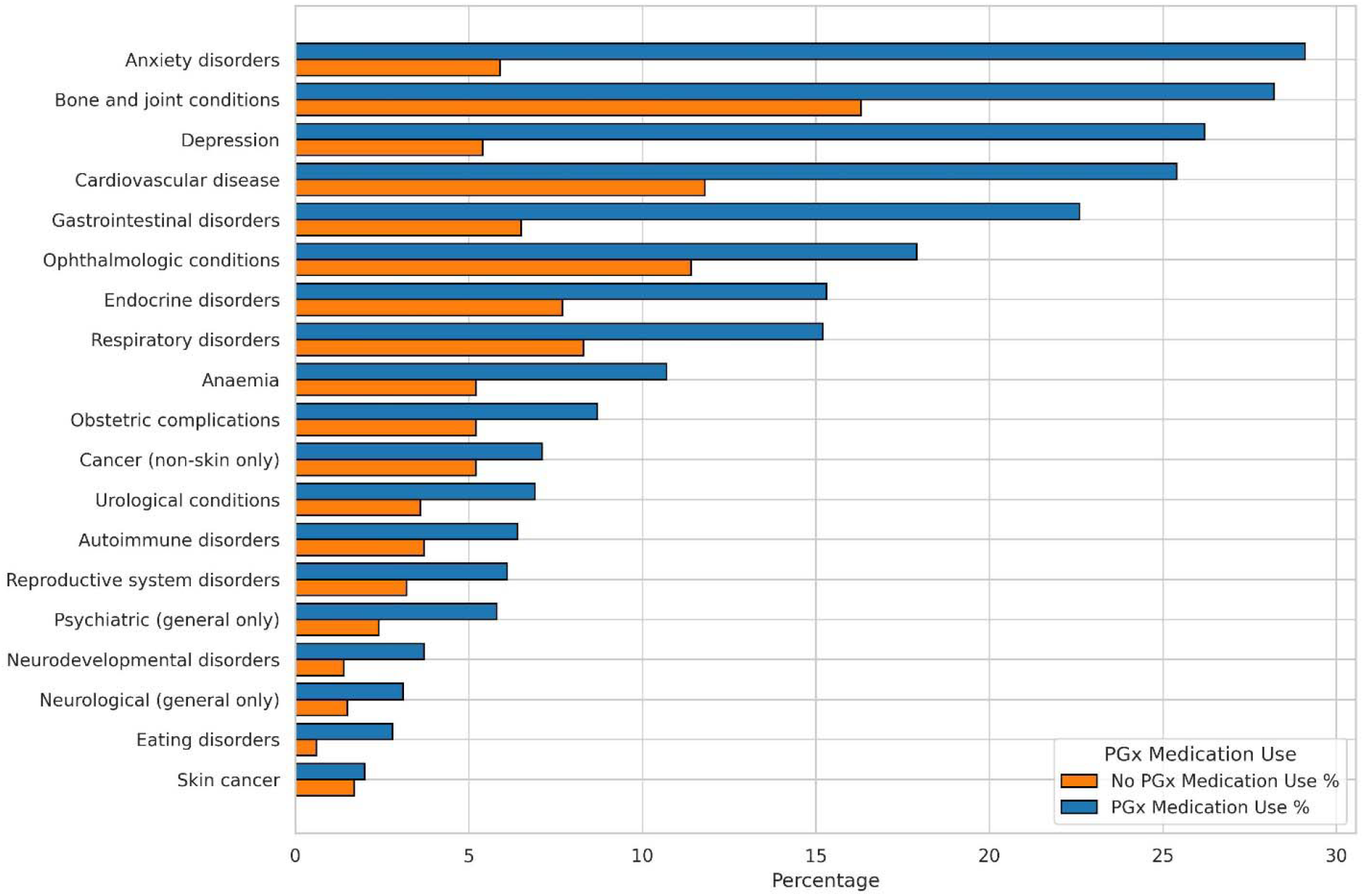
Self-reported diagnoses, stratified by self-reported PGx medication use Note: This data are mutually exclusive categories based on self-report of ever having received a diagnosis.

The ranking by prevalence of these conditions broadly corresponds to the ranking of long-term conditions reported in representative population-level resources such as the Health Survey for England (noting that OFH is UK-wide) (13), the 2022 release of which reported the five most prevalent conditions as musculoskeletal, mental, circulatory, diabetes/endocrinal metabolic, and respiratory.

Participants were more likely to self-report having received a diagnosis at older age groups, if they were female, and if they were taking a PGx medication (Figure 4).

**Figure 4.**
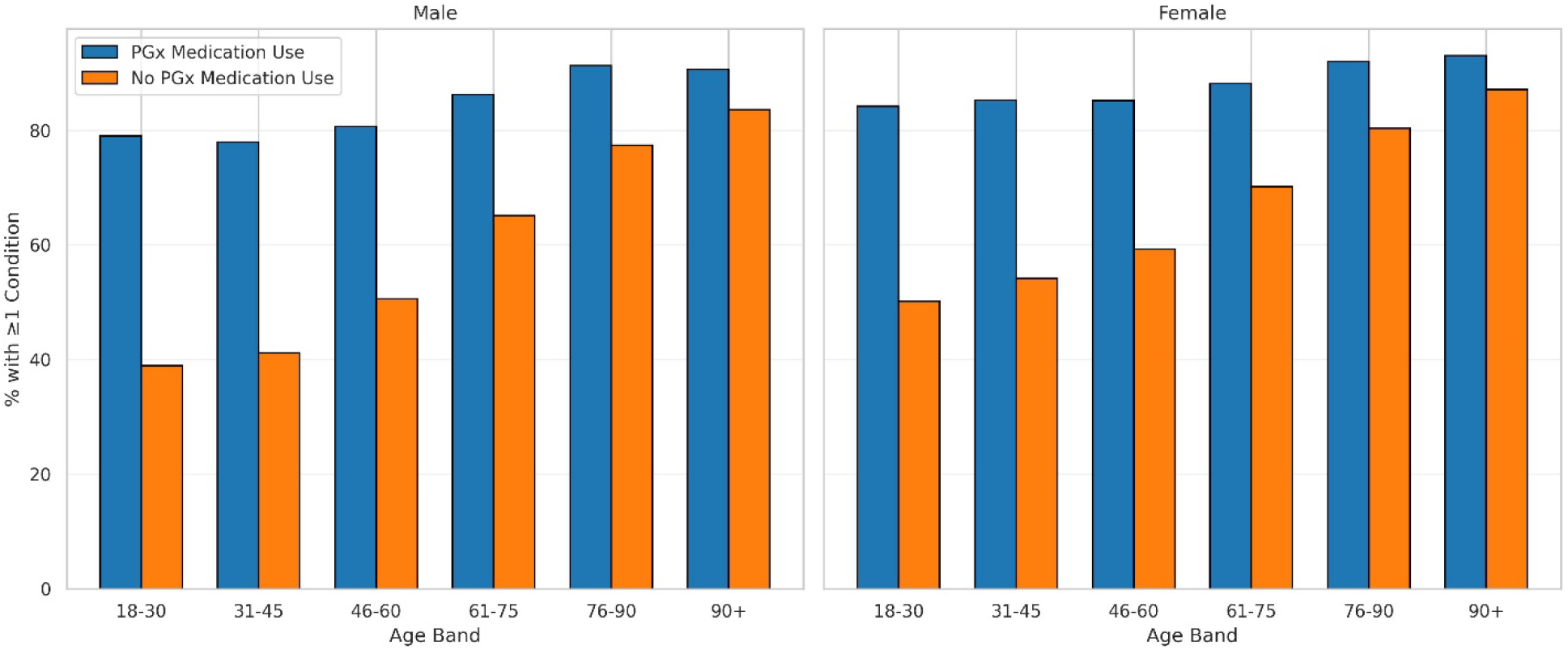
Prevalence of at least one condition by age, sex, and stratified by PGx medication use

### PGx medication use

We considered PGx medication use in the 25.2% (N=449,641) of the cohort reporting use of named drugs. PGx medication/medication groups use differed by sex in the number of reported medication groups used (Figure 5), and also in the composition of those medication groups (Figure 6).

**Figure 5.**
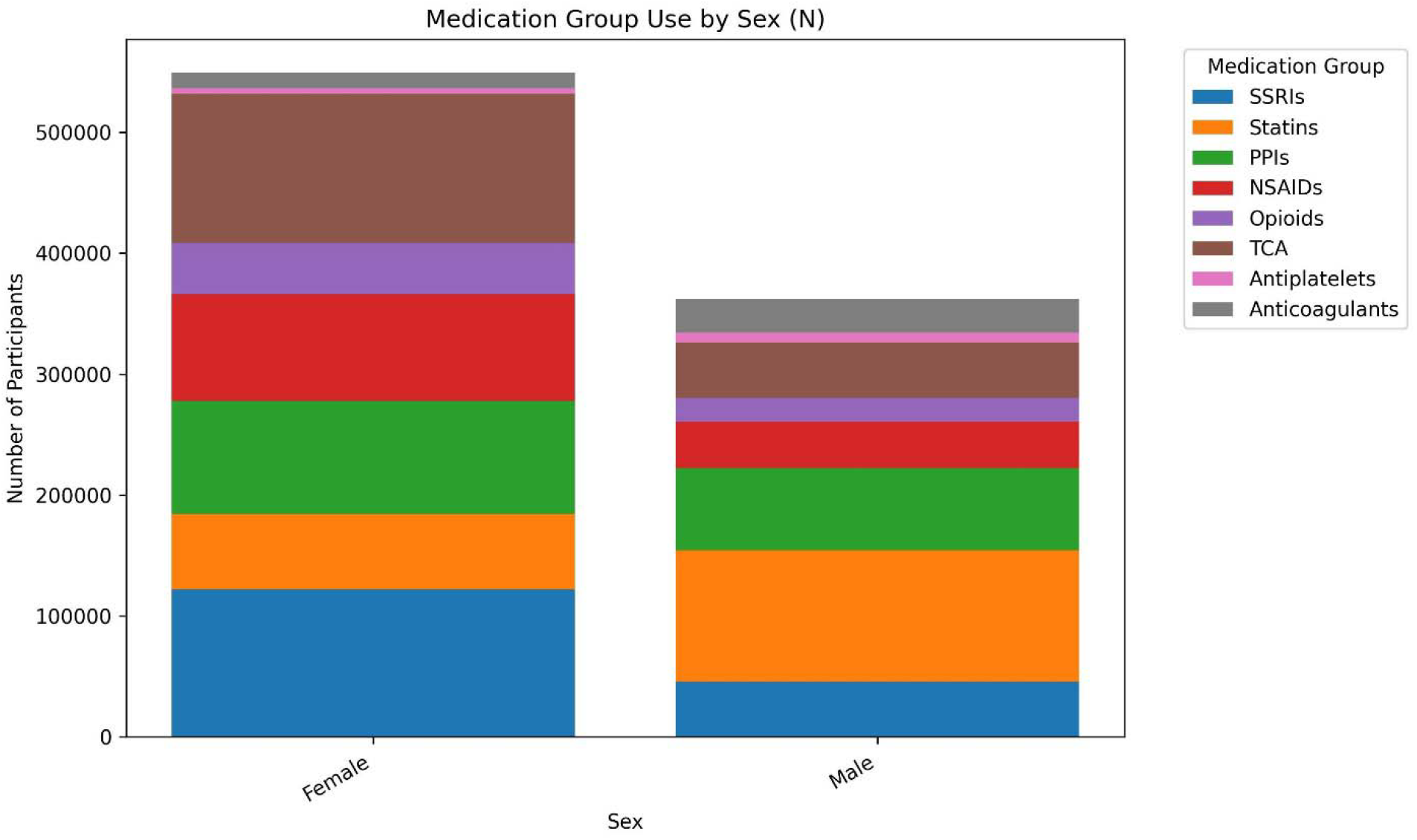
Number of patients who reported PGx use by sex and medication group

**Figure 6.**
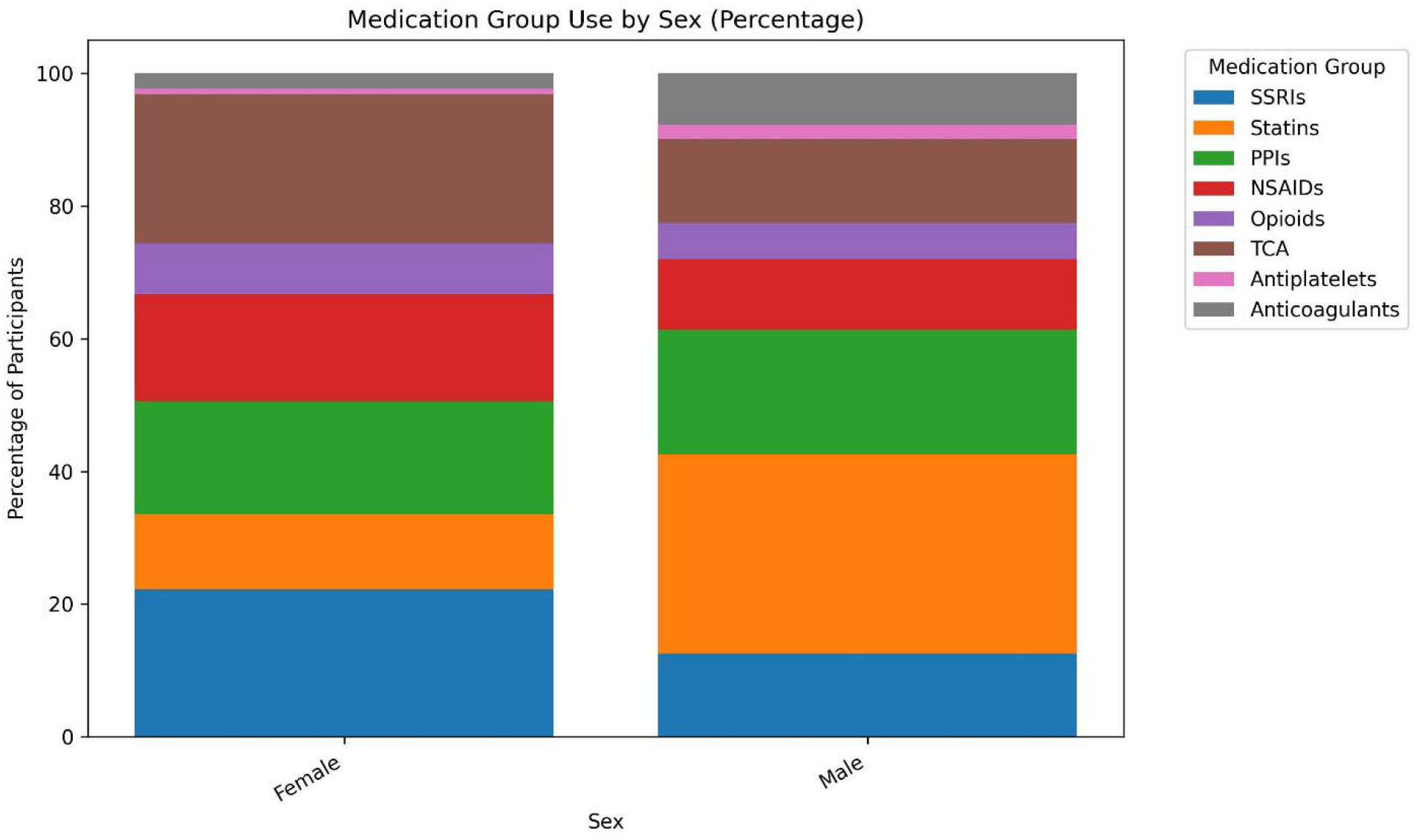
PGx medication group use by sex (as percentage shares)

Females had higher absolute use across most categories, with use of SSRIs and TCAs more widely reported among females. In contrast, males showed relatively higher use of cardiometabolic medications such as statins, antiplatelets, and anticoagulants. In this context, it is worth noting that the baseline questionnaire requested responses on regular use of drugs used “most days of the week for the last four weeks”, in which context the reported use of NSAIDs is notable.

PGx medication/medication group use also indicated notable differences by age band in terms of numbers of participants (Figure 7) and medication group mix (Figure 8).

**Figure 7.**
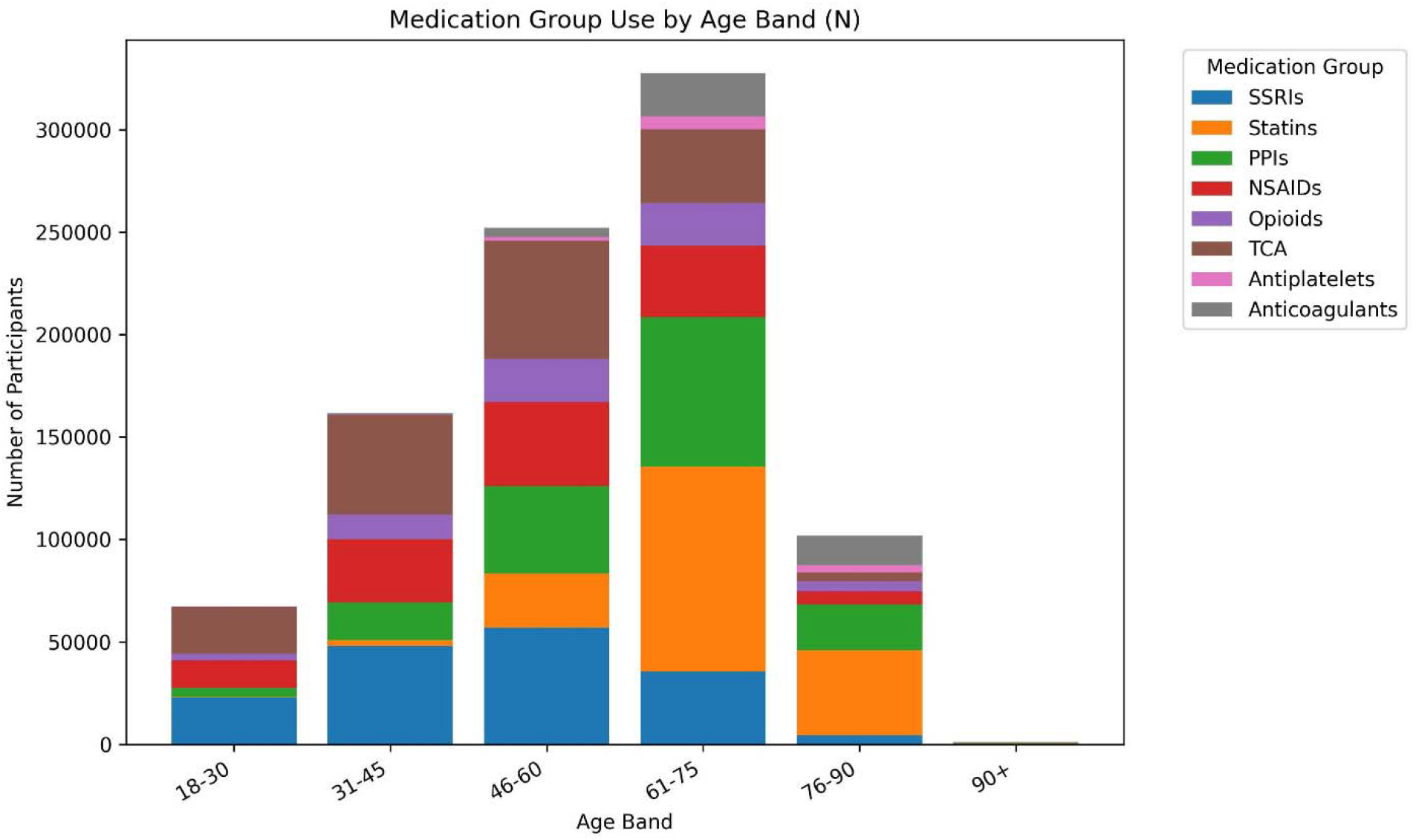
Number of participants who reported PGx use by age and medication group

**Figure 8.**
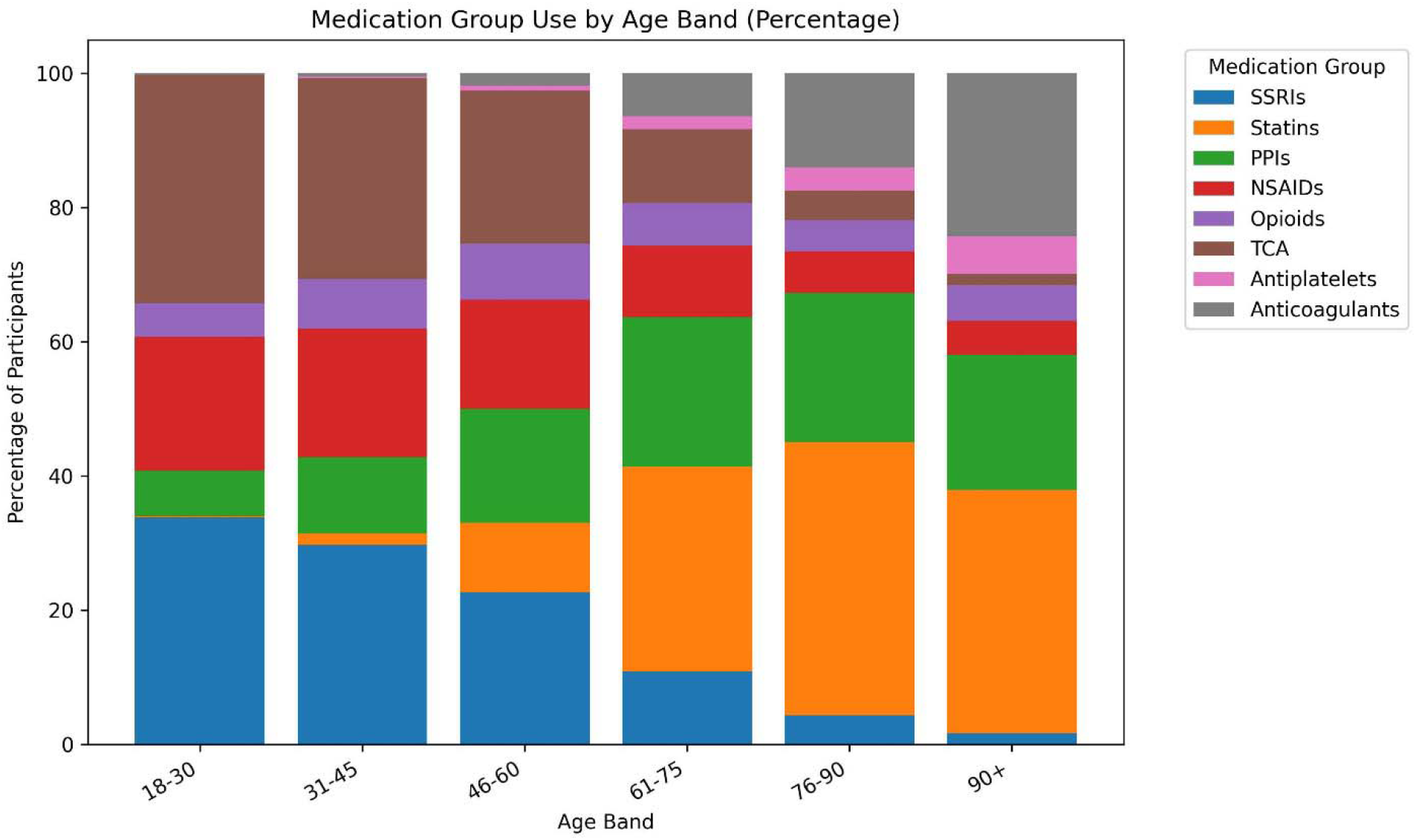
PGx medication group use by age band (as percentage shares)

PGx medication use and the number of medications used both increased with age, although with important differences in the composition of medication use. Medications related to anxiety and depression were more often reported by younger cohort participants, while cardiometabolic and gastrointestinal drugs are relatively more important for older cohorts. There was fairly consistent use of opioids over age groups, but NSAID use was more common in younger individuals.

Given that the OFH cohort is over 91% White, we report only the percentage shares rather than the absolute count of drug use by ethnicity (Figure 9).

**Figure 9.**
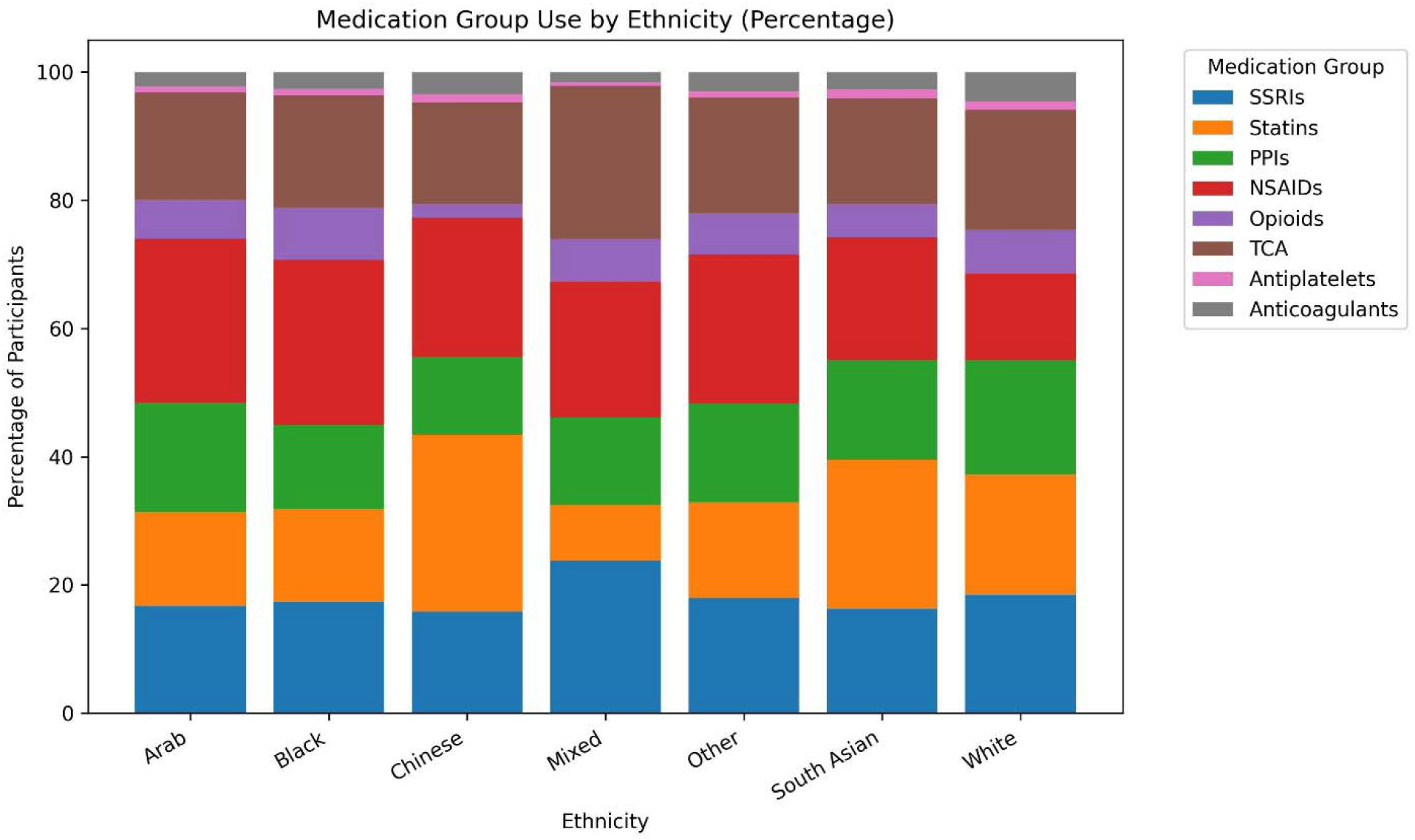
PGx medication/medication group use by ethnicity (as percentage shares)

There was slightly higher reported SSRI and TCA use by Mixed and White participants. Chinese and South Asian participants reported a high share of statin use, the latter probably reflecting higher cardiometabolic risk (14, 15) in those populations.

Finally, we consider the use of multiple drug groups. Most people used either one (42.4%) or two (39.8%) drug groups, together accounting for over 82% of users. A smaller proportion used three groups (12.2%), while very few used four or more groups. On average, individuals used 1.8 drug groups, with a median of two.

Table 2 presents the top 10 most common concurrent dual medication group combinations identified in our PGx analysis, based on absolute usage volume. For each combination, the associated pharmacogenes are also listed. Note also that we describe this as concurrent use as this is the way the baseline questionnaire requested responses from participants, but it is possible some responses do not necessarily reflect ongoing concurrent prescriptions of these drugs.

**Table 2.**
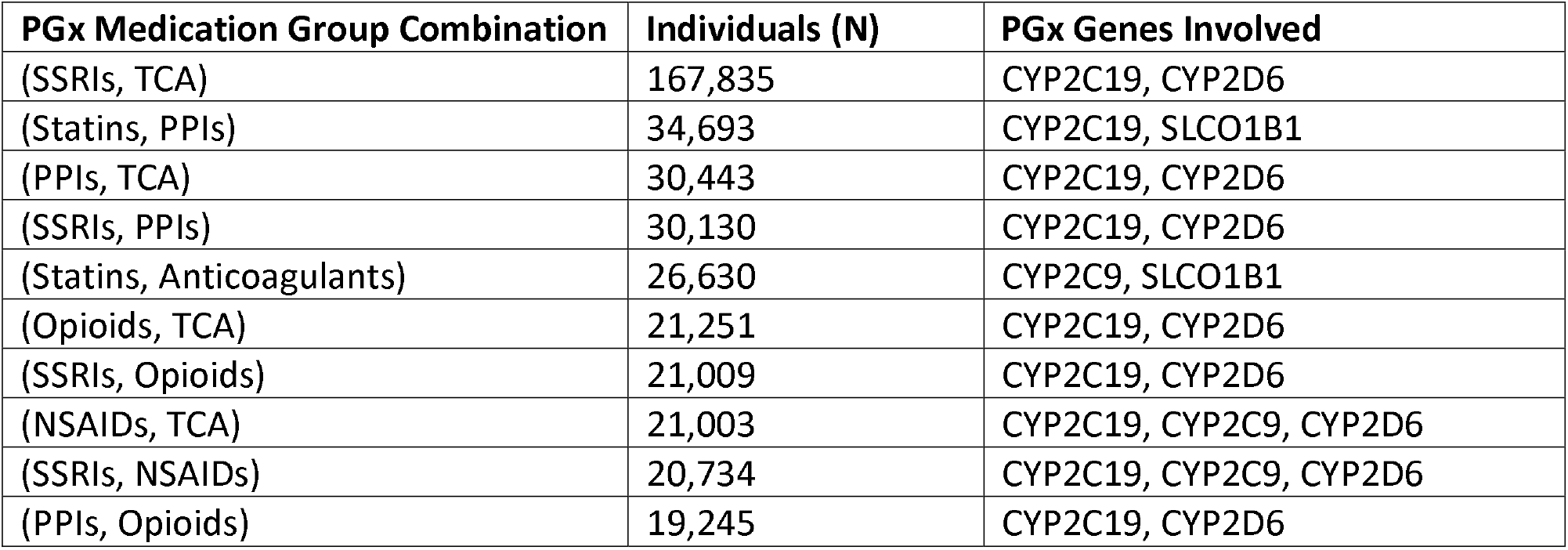
Concurrent medication/medication group use

Table 2 shows that the concurrent use of PGx drug groups influenced by different genes is common, with important roles for CYP2C19 and CYP2D6. The most common concurrent combination, reported by approximately 37% of those reporting any PGx use, involves SSRIs and TCAs. This instance of concurrent use is notable as both dose reductions and dose increases of TCAs may be required when used in combination with CYP2D6 inhibitors, such as paroxetine, fluoxetine or sertraline (16). Given that these drugs would not usually be prescribed at the same time, it is possible that this particular combination may reflect some recall bias, which can be assessed once linked prescribing data is made available for the cohort. In relation to PPIs and SSRIs (the fourth most common combination), PPIs can inhibit CYP2C19 and may lead to phenoconversion from, for example, a normal metabolizer phenotype to a “poor metabolizer” phenotype for the SSRIs (17). Some of the combinations involve 3 of the 4 genes included in our analysis, again indicating the potential complexity of the metabolic pathways involved.

## DISCUSSION

Self-reported use of 18 CPIC Level A pharmacogenetic medications, linked to four key genes, was assessed using baseline data from the OFH cohort as of June 2025. Results were stratified by age, sex, and self-reported ethnicity. Overall, 25.2% of participants reported using at least one PGx medication group. Among these participants, the number and combination of medication groups varied across demographic groups, with 37% reporting concurrent use of multiple PGx medication groups.

### Comparisons with related work

Kimpton et al (10) analysed prescribing patterns of pharmacogenetic drugs from 1993 to 2017 for a sample of 648,141 NHS patients from the Clinical Practice Research Datalink Gold data resource. During 2011–12, 58% of patients were prescribed at least one pharmacogenetic drug. The likelihood of exposure increased with age, reaching 89% for patients aged 70 years or above over the previous two decades. This study differed from the present work a number of ways, most importantly in having access to prescribing records on specific medications rather than self-report on medication classes or groups that is available in the most recent OFH data release. Kimpton et al also considered 19 pharmacogenes rather than the four considered here, which permitted identification of 63 associated drugs, compared to the 18 specific named drugs included in our analysis. Increasing use by age and differential use by sex is apparent in Kimpton et al and the present study.

Youssef et al (4) examined the impact of a hypothetical population-level pharmacogenetic screening program for nine genes related to 56 drugs. Using data on 27 million prescriptions in 2019, the authors concluded 20-25% of new prescriptions for these drugs would necessitate therapeutic intervention. McInnes et al (5) analyzed 487,409 participants in the UK Biobank and found that for 14 genes, almost 24% of individuals had been prescribed drugs likely to have an atypical pharmacogenetic response. The most frequently prescribed drugs in this category were ibuprofen, simvastatin, amitriptyline, codeine, and citalopram, all of which were mentioned by OFH participants and included in our analysis.

### Strengths and limitations

Our findings provide evidence to inform ongoing debates around the implementation of population-scale pharmacogenetic testing in the NHS and other health systems. This is the first study to assess medication use where pharmacogenetic guidance, if available, may have been used to inform prescribing decisions within the Our Future Health cohort—an emerging research resource that, like UK Biobank before it (18), is poised to play a central role in health research over the coming decades. The study lays the groundwork for future studies in OFH. In particular, when further patient-level data are released, it will be possible not only to measure the scale of such prescriptions directly, but also the accuracy of self-report that we rely on in this study. The study is also relevant to increasing emphases on preventative care, as evidenced in, for example, the NHS’s recent 10 year health plan for England (7), which makes the case for three major changes to the NHS: bringing more care from hospital settings to the community, increasing use of digital technologies, and earlier preventative interventions.

We have also developed a simple and reproducible pipeline for analyzing self-reported medication responses. Over time, this will be integrated into a broader analytical framework to support planned studies exploring associations between genetic data, PGx phenotypes, and electronic health record data. As noted, the planned linkage to primary care prescribing records will be essential to validate and refine these initial estimates, and also to enable more precise identification of PGx exposures, durations, and therapeutic indications.

However, there are several limitations to our work. The most significant is the reliance on self-reported medication use, which may be subject to recall bias or inaccuracies. Additionally, the nature of the grouped elicitation responses under broad categories (e.g., “SSRI”) without identifying specific medications, which prevents precise mapping of gene–drug interactions at the individual drug level, creates two types of bias. The first is that the results will overstate exposures for individuals using medications named in the OFH data but that don’t otherwise meet our inclusion criteria. For example, omeprazole is explicitly named in the “Proton pump inhibitors” group: “Proton pump inhibitors (e.g., omeprazole, esomeprazole, lansoprazole, rabeprazole, pantoprazole, dexlansoprazole)”. Omeprazole’s metabolism is influenced by CYP2C19 and it is supported by the highest level of evidence in CPIC guidelines. In contrast, rabeprazole does not meet this evidence threshold and would not qualify for inclusion under our criteria. However, due to the structure of these data, we cannot determine whether the participant used omeprazole or rabeprazole, but will nonetheless be included as an exposure. On the other hand, the second type of bias affecting our analysis of self-reported use of medications in classes means we are underestimating exposure to the 16 drugs that meet our analysis criteria but which aren’t explicitly named.

Additionally, our analysis focused only on four well-characterised and common pharmacogenes—*CYP2C19, CYP2C9, CYP2D6*, and *SLCO1B1*—meaning that other clinically significant gene–drug interactions, such as those involving the HLA region, were not captured in this study. However, HLA associations are generally rare and typically associated with drug hypersensitivity, and were probably less likely to be reported by participants given that the baseline questionnaire requested responses on medications taken regularly “on most days of the week for the last four weeks”. Similarly, warfarin was included because of its association with *CYP2C9*, but variants in the *VKORC1* gene are important contributors to variation in warfarin dosing (19, 20).

We did not assess dose, duration of medication exposure, or measure adverse drug reactions— all of which are likely to be important in future research (21), particularly in the context of long-term prescribing and polypharmacy. This will be facilitated by the future planned OFH data linkages with patient-level prescribing records. We also did not consider non-prescription medications that can have pharmacogenetic effects, such as St John’s Wort.

It is important to note that OFH is a volunteer-based cohort, which may differ in important ways from the general UK population. Simple, non-quantitative comparisons with English (but not UK) population norms were made. This did not find evidence of drastic differences, but this search wasn’t comprehensive and these comparisons will merit further investigation, including as the cohort expands with continued recruitment. In particular, experience with, for example, the UK Biobank cohort identified a “healthy volunteer” effect (22). This effect may be generally present in OFH, particularly so in the initial cohort of volunteers who joined in the earliest stages.

Overall, the analysis and results on their own offer some initial evidence on the possible scale of pharmacogenetic exposures in the UK population, which has implications for the wider use of PGx testing (2, 6). Further analysis of the OFH cohort, including secondary care data and particularly linked prescribing data, will further clarify this initial evidence.

## CONCLUSIONS

Use of pharmacogenetic PGx medications/medication groups associated with just four genes— *CYP2C19, CYP2D6, SLCO1B1*, and *CYP2C9* – is common across the Our Future Health (OFH) cohort and increases with age. Even among younger adults, use of PGx medications (particularly anti-depressants and analgesics) appears to be prevalent, highlighting the pervasive and persistent nature of such use throughout adulthood. Concurrent use was also frequently encountered, raising the possibility of contraindications and drug–drug/drug-gene interactions. The OFH cohort offers a unique resource for understanding self-reported PGx drug use. These findings lay groundwork for future research into pharmacogenetic implementation. The utility of the OFH cohort will be significantly enhanced through future linkage with primary care prescribing data, and richer genetic information.

## Data Availability

Access to the June 2025 data release was facilitated by the Our Future Health Early Adopters Research Scholar programme in which authors Dixon and Wright Drakesmith participated. The data were analysed in the Our Future Health DNAnexus Trusted Research Environment (https://ourfuturehealth.dnanexus.com/). Access to the data used in our analysis will be facilitated to qualifying researchers on application to Our Future Health.

## ACKNOWLEDGEMENTS

This study makes use of de-identified data held by Our Future Health. We would like to acknowledge all the research participants who have donated their data to the Our Future Health research programme. The interpretation of the information supplied by Our Future Health is that of the authors alone.

## OFH PROJECT APPROVAL

This study was approved under Project ID OFHS240243.

