## Supplementary Material for "Self-reported pharmacogenetic medication use in the *Our Future Health* cohort"

S1	CPIC LEVELS OF EVIDENCE AND DRUG SELECTION

CPIC grades (see https://cpicpgx.org/levels-of-evidence/) the strength of evidence linking genotype to phenotype as high (“Evidence includes consistent results from well-designed, well-conducted studies”, moderate (“Evidence is sufficient to determine effects, but the strength of the evidence is limited by the number, quality or consistency of the individual studies, generalizability to routine practice, or indirect nature of the evidence”), or weak (“Evidence is insufficient to assess the effects on health outcomes because of limited number or power of studies, important flaws in their design or conduct, gaps in the chain of evidence, or lack of information”).

CPIC assigns levels (A–D) to gene–drug pairs based on evidence from PharmGKB (clinical annotations or FDA label classifications of “actionable pgx”, “genetic testing recommended”, or “genetic testing required”) or through nomination. These levels are initially provisional and become final only after detailed review through CPIC guideline development. Only CPIC Level A and B pairs have sufficient evidence to support clinical prescribing recommendations.

We identified the drugs in the tables below, deduplicated the list of drugs, and then searched explicit mention of drugs/medications in questionnaire responses.

Table S1	CYP2C19 drug guidelines

| **Gene** | **Drug** | **Guideline** | **CPIC Level** | **Level Status** |
| --- | --- | --- | --- | --- |
| CYP2C19 | Amitriptyline | <https://cpicpgx.org/guidelines/guideline-for-tricyclic-antidepressants-and-cyp2d6-and-cyp2c19/> | A | Final |
| CYP2C19 | Citalopram | <https://cpicpgx.org/guidelines/cpic-guideline-for-ssri-and-snri-antidepressants/> | A | Final |
| CYP2C19 | Clopidogrel | <https://cpicpgx.org/guidelines/guideline-for-clopidogrel-and-cyp2c19/> | A | Final |
| CYP2C19 | Escitalopram | <https://cpicpgx.org/guidelines/cpic-guideline-for-ssri-and-snri-antidepressants/> | A | Final |
| CYP2C19 | Lansoprazole | <https://cpicpgx.org/guidelines/cpic-guideline-for-proton-pump-inhibitors-and-cyp2c19/> | A | Final |
| CYP2C19 | Omeprazole | <https://cpicpgx.org/guidelines/cpic-guideline-for-proton-pump-inhibitors-and-cyp2c19/> | A | Final |
| CYP2C19 | Pantoprazole | <https://cpicpgx.org/guidelines/cpic-guideline-for-proton-pump-inhibitors-and-cyp2c19/> | A | Final |
| CYP2C19 | Sertraline | <https://cpicpgx.org/guidelines/cpic-guideline-for-ssri-and-snri-antidepressants/> | A | Final |
| CYP2C19 | Voriconazole | <https://cpicpgx.org/guidelines/guideline-for-voriconazole-and-cyp2c19/> | A | Final |

Table S2	CYP2C9 drug guidelines

| **Gene** | **Drug** | **Guideline** | **CPIC Level** | **Level Status** |
| --- | --- | --- | --- | --- |
| CYP2C9 | Celecoxib | <https://cpicpgx.org/guidelines/cpic-guideline-for-nsaids-based-on-cyp2c9-genotype/> | A | Final |
| CYP2C9 | Flurbiprofen | <https://cpicpgx.org/guidelines/cpic-guideline-for-nsaids-based-on-cyp2c9-genotype/> | A | Final |
| CYP2C9 | Fluvastatin | <https://cpicpgx.org/guidelines/cpic-guideline-for-statins/> | A | Final |
| CYP2C9 | Ibuprofen | <https://cpicpgx.org/guidelines/cpic-guideline-for-nsaids-based-on-cyp2c9-genotype/> | A | Final |
| CYP2C9 | Lornoxicam | <https://cpicpgx.org/guidelines/cpic-guideline-for-nsaids-based-on-cyp2c9-genotype/> | A | Final |
| CYP2C9 | Meloxicam | <https://cpicpgx.org/guidelines/cpic-guideline-for-nsaids-based-on-cyp2c9-genotype/> | A | Final |
| CYP2C9 | Phenytoin | <https://cpicpgx.org/guidelines/guideline-for-phenytoin-and-cyp2c9-and-hla-b/> | A | Final |
| CYP2C9 | Piroxicam | <https://cpicpgx.org/guidelines/cpic-guideline-for-nsaids-based-on-cyp2c9-genotype/> | A | Final |
| CYP2C9 | Tenoxicam | <https://cpicpgx.org/guidelines/cpic-guideline-for-nsaids-based-on-cyp2c9-genotype/> | A | Final |
| CYP2C9 | Warfarin | <https://cpicpgx.org/guidelines/guideline-for-warfarin-and-cyp2c9-and-vkorc1/> | A | Final |

Table S3	CYP2D6 drug guidelines

| **Gene** | **Drug** | **Guideline** | **CPIC Level** | **Level Status** |
| --- | --- | --- | --- | --- |
| CYP2D6 | Amitriptyline | <https://cpicpgx.org/guidelines/guideline-for-tricyclic-antidepressants-and-cyp2d6-and-cyp2c19/> | A | Final |
| CYP2D6 | Atomoxetine | <https://cpicpgx.org/guidelines/cpic-guideline-for-atomoxetine-based-on-cyp2d6-genotype/> | A | Final |
| CYP2D6 | Codeine | <https://cpicpgx.org/guidelines/guideline-for-codeine-and-cyp2d6/> | A | Final |
| CYP2D6 | Nortriptyline | <https://cpicpgx.org/guidelines/guideline-for-tricyclic-antidepressants-and-cyp2d6-and-cyp2c19/> | A | Final |
| CYP2D6 | Ondansetron | <https://cpicpgx.org/guidelines/guideline-for-ondansetron-and-tropisetron-and-cyp2d6-genotype/> | A | Final |
| CYP2D6 | Paroxetine | <https://cpicpgx.org/guidelines/cpic-guideline-for-ssri-and-snri-antidepressants/> | A | Final |
| CYP2D6 | Tamoxifen | <https://cpicpgx.org/guidelines/cpic-guideline-for-tamoxifen-based-on-cyp2d6-genotype/> | A | Final |
| CYP2D6 | Tramadol | <https://cpicpgx.org/guidelines/guideline-for-codeine-and-cyp2d6/> | A | Final |
| CYP2D6 | Tropisetron | <https://cpicpgx.org/guidelines/guideline-for-ondansetron-and-tropisetron-and-cyp2d6-genotype/> | A | Final |
| CYP2D6 | Vortioxetine | <https://cpicpgx.org/guidelines/cpic-guideline-for-ssri-and-snri-antidepressants/> | A | Final |

Table S4	SLCO1B1 drug guidelines

| **Gene** | **Drug** | **Guideline** | **CPIC Level** | **Level Status** |
| --- | --- | --- | --- | --- |
| SLCO1B1 | Atorvastatin | <https://cpicpgx.org/guidelines/cpic-guideline-for-statins/> | A | Final |
| SLCO1B1 | Fluvastatin | <https://cpicpgx.org/guidelines/cpic-guideline-for-statins/> | A | Final |
| SLCO1B1 | Lovastatin | <https://cpicpgx.org/guidelines/cpic-guideline-for-statins/> | A | Final |
| SLCO1B1 | Pitavastatin | <https://cpicpgx.org/guidelines/cpic-guideline-for-statins/> | A | Final |
| SLCO1B1 | Pravastatin | <https://cpicpgx.org/guidelines/cpic-guideline-for-statins/> | A | Final |
| SLCO1B1 | Rosuvastatin | <https://cpicpgx.org/guidelines/cpic-guideline-for-statins/> | A | Final |
| SLCO1B1 | Simvastatin | <https://cpicpgx.org/guidelines/cpic-guideline-for-statins/> | A | Final |

S2	SPECIFIC QUESTIONS ASKED ABOUT MEDICATION USAGE

The baseline questionnaire (version 2) posed questions as follows for medications potentially eligible for inclusion in our analysis.

Antidepressants

- “Antidepressant (e.g. fluoxetine, amitriptyline, sertraline, mirtazapine)” under an “antidepressant medication” category
- “Selective serotonin reuptake inhibitor (SSRI, e.g., Fluoxetine, citalopram, escitalopram, paroxetine, sertraline)
- Tricyclic antidepressants (e.g., amitriptyline, clomipramine)
- Other (e.g., mirtazapine, venlafaxine, duloxetine)”
- “Amitriptyline for migraines” under a neurological category

Statins

- “Statins” under “medication for diabetic health?”
- “Cholesterol lowering medication/statins” under “medication for heart or circulatory disorders”

Proton pump inhibitors

- “Proton pump inhibitors (e.g., omeprazole, esomeprazole, lansoprazole, rabeprazole, pantoprazole, dexlansoprazole)” under “medication [sic] digestive problems, acid reflux or liver problems?”

NSAIDs

- “Ibuprofen (e.g., Neurofen)” under a pain category

Opioids

- “Opioids (e.g., codeine, tramadol, morphine, fentanyl, oxycodone, buprenorphine diamorphine)” under the same pain category as ibuprofen

Anticoagulants

- “Anticoagulant (blood thinners, e.g., warfarin, Rivaroxaban, dabigatran, apixaban and edoxaban)” under “medication for heart or circulatory disorders”

Antiplatelets

- “Anti-platelet (Clopidogrel)” under “medication for heart or circulatory disorders”

S3 MEDICATIONS NOT MEETING INCLUSION CRITERIA

Medications meeting CPIC criteria but not explicitly mentioned in the baseline questionnaire and excluded from analysis: atomoxetine, celecoxib, flurbiprofen, lornoxicam, lovastatin, meloxicam, nortriptyline, ondansetron, phenytoin, piroxicam, pitavastatin, tamoxifen, tenoxicam, tropisetron, voriconazole, vortioxetine.
